# A systematic review of behavioral interventions to improve maternal outcomes for women in the United States at high risk for adverse pregnancy outcomes

**DOI:** 10.1101/2025.07.18.25331742

**Authors:** Jennifer E. Phipps, Indira D’Souza, Nikita Satish, Audriana Ketchersid, Mackenzie D.M. Whipps, Megan Van Noord, Imo Ebong, Tanya Khemet Taiwo, Leanna S. Sudhof, Victoria Keeton, Herman L. Hedriana, Leigh Ann Simmons

## Abstract

**Introduction:** Adverse pregnancy outcomes (APOs), including hypertensive disorders, gestational diabetes, and preterm birth, affect 10–20% of U.S. pregnancies and substantially increase long-term cardiovascular disease (CVD) risk. While behavioral interventions during pregnancy may reduce risk factors for APOs, evidence remains limited on which strategies are effective for high-risk women. This systematic review addresses this gap by examining patterns in intervention design, outcome reporting, and areas where research fails to meet the needs of at-risk pregnant populations.

**Methods:** We searched PubMed and the Cochrane Central Register of Controlled Trials for articles published from January 1, 2005, to June 30, 2024. Following PRISMA guidelines, we included U.S.-based randomized controlled trials of behavioral interventions: diet, physical activity, sleep, mindfulness, or self-monitoring, delivered during pregnancy to women at high risk of APOs. See Supplemental Figure S4 for the PRISMA Checklist. Two reviewers independently performed screening, data extraction, and risk-of-bias assessment using the Cochrane Risk of Bias Tool.

**Results:** Of 3,261 studies screened, 43 met inclusion criteria. Most interventions focused on individuals with prepregnancy overweight or obesity, often excluding those with other CVD risk factors. Thirty-six trials reported improvements in outcomes such as gestational weight gain (GWG), postpartum weight retention, blood pressure, or biomarkers. Interventions combining diet and physical activity showed the greatest benefit. However, only four trials demonstrated improvements in clinical outcomes like gestational diabetes, hypertensive disorders, or lipid profiles. Outcome measures and timelines varied widely, and few trials included postpartum follow-up or assessed outcomes beyond GWG.

**Discussion:** Although behavioral interventions during pregnancy are growing in number, trials are limited by heterogeneous designs, narrow inclusion criteria, and inconsistent outcome reporting. The focus on GWG and exclusion of women with complex risk profiles reduce generalizability. Inclusive research with standardized outcomes is urgently needed to improve maternal health and reduce long-term CVD risk.

## INTRODUCTION

Approximately 10-20% of pregnant women in the United States (US) will experience an adverse pregnancy outcome (APO) such as pre-eclampsia or other hypertensive disorders of pregnancy (HDPs), gestational diabetes mellitus (GDM), infant growth issues, or pre-term birth. APOs are associated with significant morbidity and mortality, especially for Black and African American women who experience additional social and health inequities that increase their risk for pregnancy complications.^1, 2^ They are also associated with higher risk for cardiovascular disease (CVD) both in the short- and long-term.^3^ HDPs were reported in up to 15.9% of pregnancies in the US in 2019, up from 13% in 2017, with rates highest in non-Hispanic Black and African American women.^1^ Rates of GDM are also increasing, affecting approximately 8% of pregnancies in the US in 2021, with rates highest in Asian women.^4^ Pre-pregnancy obesity and other pre-existing CVD risk factors (e.g. hypertension, diabetes) increase the likelihood of developing APOs and are becoming more prevalent in women of childbearing age in the US.^5^

Attempts to reduce APOs have been tested in the US using various behavioral change strategies. Interventions have focused on improving diet, reducing gestational weight gain (GWG), increasing physical activity, lowering stress through mindfulness, or a combination of these.^6–9^ However, many of these interventions were designed for pregnant women in general, often excluding specific high-risk conditions such as those with pre-existing CVD risk factors due to potential complications these participants may develop.^10^ Even among interventions trialed for low-risk pregnant women, findings have been mixed. Lifestyle changes may be beneficial in achieving recommended GWG,^8, 11^ particularly for pregnant women who enter pregnancy with normal BMI.^12^ Interventions to reduce GDM rates have shown mixed results. While those that combined dietary and physical activity interventions showed the most promise, they lacked substantial evidence.^13^ Lifestyle interventions targeting HDP rates have been generally unsuccessful.^10, 14^ Interventions that included physical activity were most likely to lower blood pressure.^9^ Given the mixed results among low-risk pregnancies, it is crucial to investigate if behavioral change strategies may have better outcomes in high-risk pregnant women who have increased likelihood of poor pregnancy outcomes.

The aim of this systematic review was to fill a gap in the literature by identifying, evaluating, and summarizing findings from randomized controlled trials of behavioral interventions designed for pregnant women at high risk for APOs. The purpose of the study was to identify the most beneficial interventions for specific outcomes and evaluate the presence of gaps in the literature on behavioral interventions for this patient population. These findings will inform how midwives and other health care providers can advance practice and research strategies to improve maternal health for these high-risk groups in the US.

## METHODS

Detailed methods can be found in the preregistered published protocol.^15^ The extracted data may be made available with reasonable requests. Institutional review board approval was not required.

### Search strategy

Extensive literature searches were conducted in December 2021 and April 2023 in Ovid MEDLINE (via PubMed) and the Cochrane Central Register of Controlled Trials (CENTRAL) for articles published from 2005 through the search dates. An updated search was completed on June 30, 2024. The search strategy focused on concepts related to pregnancies at high risk for adverse pregnancy outcomes (APOs) and behavioral interventions. The full search strategy is available in Supplemental Table S2.

### Inclusion and exclusion criteria

To be included in this review, studies had to meet the following criteria: 1) included pregnant participants; 2) used a parallel-arm randomized controlled trial (RCT) study design; 3) were performed in the US, to minimize the influence of differing health care systems on maternal outcomes;^16^ 4) were published between January 1, 2005 and June 30, 2024; 5) investigated a behavioral intervention (e.g., diet, physical activity, mindfulness) designed for pregnant women at high risk for APOs (e.g. pre-pregnancy obesity, previous or current diagnosis of HDP or GDM); 6) implemented the intervention during pregnancy; and 7) reported on maternal pregnancy outcomes (e.g. blood pressure, GWG, diagnosis of complications). Exclusion criteria were: 1) animal studies, 2) studies that only assessed intervention adherence, 3) interventions designed for obstetric providers, 4) crossover or cluster RCT study design; and 5) follow-up studies from previous RCTs. See the published protocol for a complete list of APO risk factors used in screening.^15^

### Screening, data extraction, and risk of bias analysis

Covidence systematic review software (Veritas Health Innovation, Melbourne, Australia) was used for screening and data extraction. Covidence’s standard template for risk of bias analysis was used to assess risk of bias; this template implements the Cochrane Risk of Bias version 1 tool.^17^ A team of five individuals (two PhD-prepared researchers and three students) carried out the review tasks. Title/abstract screening was independently performed by two reviewers (one PhD-prepared), selected from this team. Disagreements were resolved by a co-author who was not involved in the review process. The same process was used for full-text screening, which included merging any studies that reported on the same cohorts of trial participants. Data extraction was also conducted by two independent reviewers (one PhD-prepared), and risk of bias analysis was performed by a different pair of independent reviewers from the same team. A final consensus on extracted data and risk of bias ratings was conducted by a PhD-prepared reviewer. Because trials varied in how and when GWG was measured, we extracted GWG values as reported by each study, whether they reflected total pregnancy weight gain or only weight gained during the intervention period.

## RESULTS

### Included Studies

Initially, N=4,742 references were returned and 43 were included in the analysis (Figure 1).^18–60^ Ultimately, results from 5,778 pregnant women were analyzed with studies ranging in sample size from 12 to 958. A full data table is available in the Supplement (Supplemental Table S1).

**Figure 1.**
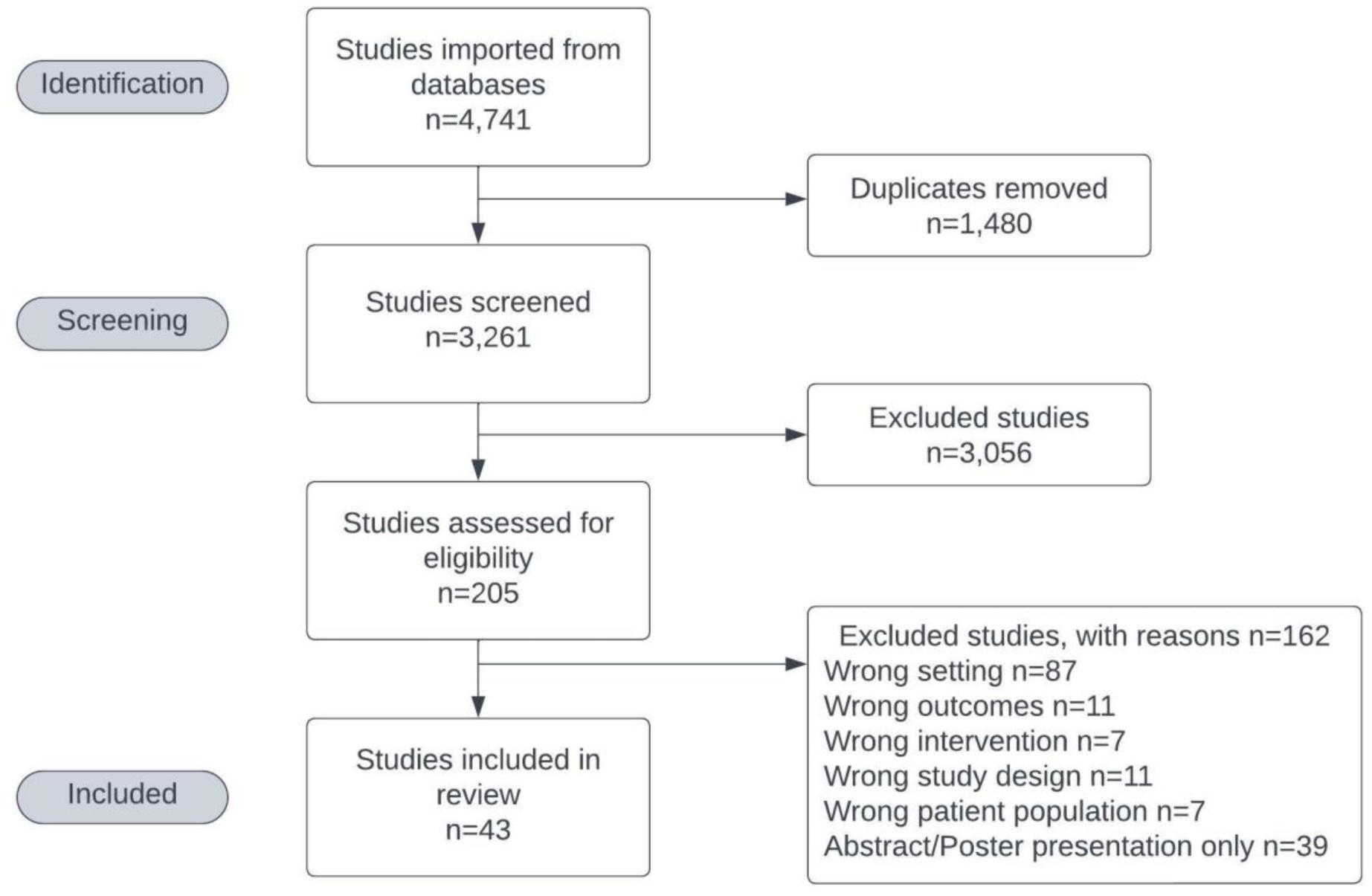
PRISMA Flowchart

### Study Characteristics

Behavioral intervention targets included diet, physical activity, mindfulness (e.g., stress management, mindfulness training), self-monitoring behaviors (e.g., self-weighing or home glucose monitoring), sleep, and health coaching or counseling regarding diet and other lifestyle behaviors (Table 1). Study participants included pregnant women with overweight or obesity; those at high risk for APOs due to family history, a prior diagnosis, or a current diagnosis; those with a pre-existing CVD risk factor such as diabetes or hypertension; or those with a combination of these factors. Recruitment for 88% of trials occurred at clinics; 12% included social media or other online recruitment methods. Maternal outcomes that were extracted included: diagnosis of an APO, blood pressure, blood biomarkers related to CVD (e.g. cholesterol, triglycerides, blood glucose), GWG, and postpartum weight retention (PPWR).

**Table 1.**

| **Table 1. Trial Characteristics** | **N** | **(%)** |
| --- | --- | --- |
| Type of Trial |  |  |
| Parallel arm RCT<sup>18-60</sup> | 43 | 100% |
| Method of Recruitment |  |  |
| Clinic/standard | 38 | 88% |
| Social media/online<sup>20, 27, 30, 50, 60</sup> | 5 | 12% |
| Behavior |  |  |
| Diet<sup>19, 21, 22, 25, 32, 41, 43, 51, 52</sup> | 9 | 21% |
| Physical Activity<sup>31, 44, 45, 50, 54</sup> | 5 | 12% |
| Diet and Physical Activity<sup>18, 26-30, 33, 36-38, 40, 47-49, 55-57</sup> | 17 | 40% |
| Glucose self-monitoring<sup>23</sup> | 1 | 2% |
| Mindfulness training<sup>59</sup> | 1 | 2% |
| Sleep<sup>58</sup> | 1 | 2% |
| Diet and Other*<sup>24, 35, 46, 53</sup> | 4 | 9% |
| Physical Activity and Other<sup>60</sup> | 1 | 2% |
| Diet, Physical Activity and Other*<sup>20, 34, 39, 42</sup> | 4 | 9% |
| Inclusion Criteria |  |  |
| Overweight/Obesity<sup>18, 20, 21, 27, 28, 30-40, 42, 47, 48, 50-52, 55</sup> | 23 | 53% |
| Overweight/Obesity and current GDM diagnosis<sup>19, 22, 43, 56</sup> | 4 | 9% |
| Current or previous GDM<sup>23, 25, 26, 29, 41, 44-46, 49, 53, 57, 58</sup> | 12 | 28% |
| Current or previous hypertensive disorder<sup>54, 59</sup> | 2 | 5% |
| Pre-existing type 1 diabetes<sup>24</sup> | 1 | 2% |
| High sedentary behavior risk and high APO risk<sup>60</sup> | 1 | 2% |
| End of intervention |  |  |
| Birth- no postpartum monitoring | 33 | 77% |
| Birth- with postpartum monitoring<sup>32, 35, 47, 48, 50, 52, 55</sup> | 7 | 16% |
| Postpartum<sup>26, 44, 49</sup> | 3 | 7% |

\*Other may include stress-management, social media groups, coaching, counseling, or self-monitoring.

### Details of study cohorts

Table 2 summarizes trial participant data. Most participants identified as White and had obesity. Race and ethnicity^60^ data were reported as separate entities in only 13 trials. Two trials allowed participants to report more than one race or ethnicity. Several trials had missing race and/or ethnicity data, and one trial reported aggregate race and ethnicity data across all participants without separating by intervention arm. As a result, the percentages and totals in Table 2 do not always add up to 100%, and the race and ethnicity rows do not match the total number of participants. On average, study participants were 30 years old, nearly half were nulliparous, and most began their intervention early in the second trimester. Of the 28 trials that used BMI as an inclusion criteria, six required participants to have obesity, 20 allowed either overweight or obesity, and two allowed a broader range—from upper-normal BMI to obesity (i.e., ≥22^32^ and 20-45^33^). Across studies that reported weight gain, participants gained an average of 20–25 pounds during pregnancy, over the course of the interventions, which all began during pregnancy and, in some cases, continued into the postpartum period. Because interventions did not always begin early in pregnancy, reported GWG values varied across studies and were not always equivalent to total gestational weight gain.

**Table 2.**
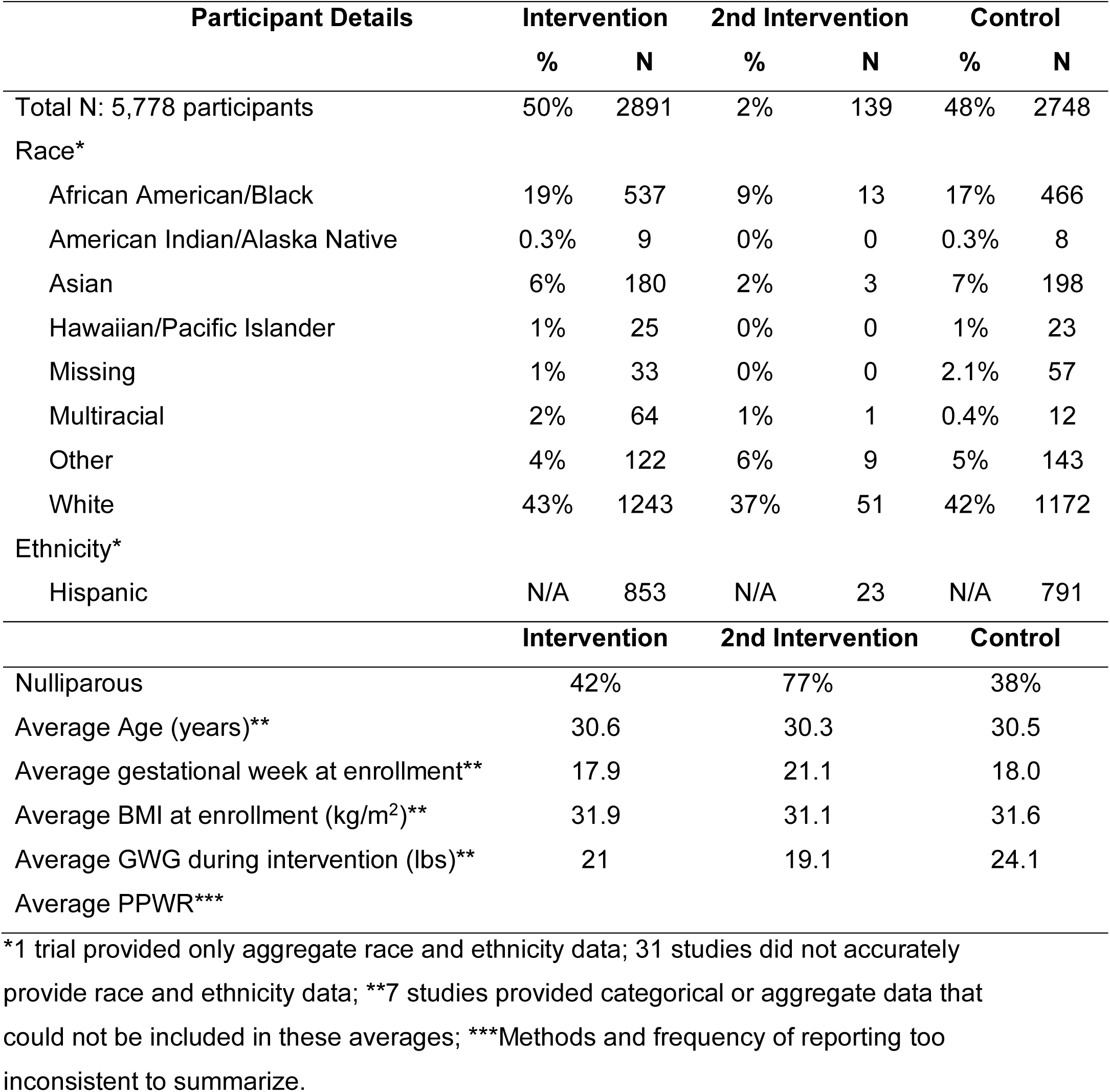
Participant Details Intervention 2nd Intervention Control.

Three interventions extended into the postpartum period^26, 44, 49^ and seven prenatal interventions continued follow-up participants into the postpartum period (ranging from 2 weeks to 1 year).^32, 35, 47, 48, 50, 52, 55^ Of the 43 interventions, 30 excluded participants with pre-existing hypertension, diabetes, or other CVD risk factors. Only two interventions were specifically designed for participants with a chronic condition (Type 1 diabetes,^24^ and chronic hypertension^59^). Among the 41 studies that reported participant location, 28 US states were represented (Supplemental Figure S1). Two multi-state studies did not specify which states were included.^25, 53^

### Bias Analysis

The greatest sources of bias in the included trials were in (1) allocation concealment, (2) sequence generation, and (3) blinding of participants, study personnel, and/or outcome assessors. Most of the sources of bias came from a lack of information in the manuscripts and available data registered with clinicaltrials.gov. Other than the lack of blinding, risk of bias overall was relatively low for all included studies (see Supplemental Figure S2 and Supplemental Figure S3).

### Success of Interventions

Of the 43 trials, 36 reported statistically significant results, while seven reported null results. The most common statistically significant outcome was reduced GWG (Table 3). Figure 2 categorizes the significant findings from Table 4 by trial type. Most studies focused on diet, physical activity, or a combination of the two. Trials that combined diet and physical activity most frequently showed significant improvements in maternal outcomes, including reduced GWG, improved physical activity levels, and reduced PPWR. In contrast, trials that focused solely on physical activity had the most null results. The seven trials with null results aimed to reduce postpartum mean fasting glucose levels through a prenatal and postpartum physical activity intervention for pregnant women with GDM,^44^ improve prenatal blood glucose levels through a dietary intervention for pregnant women with GDM,^41^ improve maternal insulin resistance and 24-hr glycemia with a combined dietary and physical activity intervention for pregnant women with GDM,^56^ reduce GDM risk through a physical activity intervention,^45^ and reduce GWG with a combined dietary and physical activity intervention.^30, 33, 35^ PPWR was significantly reduced in three trials.^26, 32, 48^

**Figure 2.**
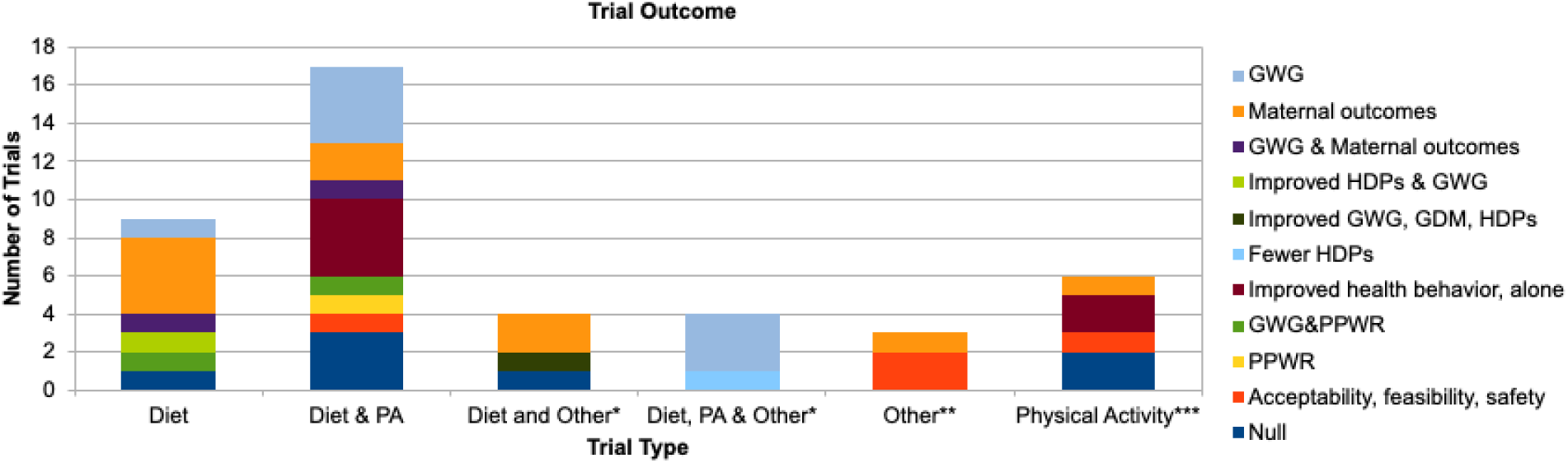
Significant trial outcomes by trial type. *Addition of ‘other’ to an intervention may include mindfulness, self-monitoring, or coaching/counseling. **’Other’ as an intervention type represents three studies that trialed the following: glucose self-monitoring, sleep, and mindfulness training. ***Physical activity also includes one study that trialed physical activity along with coaching and a social media group.

**Table 3.**

| **Table 3. Significant Outcome Measures** | **Trials (N)** |
| --- | --- |
| Reduced GWG<sup>21, 28, 34, 36, 39, 40, 42, 55</sup> | 8 |
| Reduced PPWR<sup>26</sup> | 1 |
| Reduced GWG & PPWR<sup>32, 48</sup> | 2 |
| Reduced GWG and improved maternal outcomes* <sup>22, 38</sup> | 2 |
| Improved maternal outcomes* <sup>19, 24, 25, 29, 43, 46, 51, 54, 57, 59</sup> | 10 |
| Reduced rates of HDPs<sup>20</sup> | 1 |
| Reduced rates of HDPs and lower GWG<sup>52</sup> | 1 |
| Reduced rates of GDM and HDPs, lower GWG<sup>53</sup> | 1 |
| Improved health behaviors** <sup>18, 37, 47, 49, 50, 60</sup> | 6 |
| Acceptability/feasibility only<sup>23, 27, 31, 58</sup> | 4 |
| Null<sup>30, 33, 35, 41, 44, 45, 56</sup> | 7 |

\*Maternal Outcomes included improved gut microbiome, improved blood biomarkers (triglycerides, IL-6, free fatty acids, cholesterol, C-reactive protein, A1c, glucose, insulin, or transferrin levels) or lower blood pressure. \*\*Improved health behaviors include: self-monitoring of weight,<sup>18</sup> self-monitoring of physical activity,<sup>18</sup> increased physical activity minutes,<sup>18</sup> reduced sedentary time,<sup>60</sup> improved attitudes about nutritious eating and physical activity,<sup>37</sup> increase in vigorous-intensity exercise,<sup>47</sup> increased minutes of walking,<sup>50</sup> and decreased dietary fat intake.<sup>49</sup>

## DISCUSSION

Since 2005, 43 parallel arm RCTs of behavioral interventions have targeted pregnant women at high risk for APOs.^18–60^ Most of these trials focused on overweight and obesity as risk factors. In total, 30 trials specifically <u>excluded</u> women with pre-existing conditions associated with CVD-risk and APOs; in 14 of these cases, the authors explicitly did so to isolate the effects of the intervention on the risk of HDP or GDM.^19, 22, 23, 25, 26, 41, 43–46, 49, 53, 54, 56^ However, women with a single CVD risk factor or who experience an APO are at heightened risk of developing additional CVD risk factors and even subsequent or secondary APOs.^61, 62^ Thus, excluding these women from trials limits the generalizability of the findings for those at highest risk and my omit individuals who could benefit from such interventions.

Additionally, BMI was measured in varied ways across studies, frequently self-reported, sometimes based on first trimester weight, and other times on pre-pregnancy weight. These differences in BMI assessment, along with variability in reported outcomes (e.g., some trials reported blood pressure while others did not; some included postpartum follow-up while others did not; some measured lipid levels while others excluded such measures), limited the comparability of findings. These inconsistencies hinder our ability to draw firm conclusions about which interventions effectively improve maternal outcomes. Among the trials that successfully reduced GWG, only four demonstrated improvements in maternal outcomes, specifically in GDM, HDPs, diastolic blood pressure, and blood triglyceride levels.^22, 38, 52, 53^

The heterogeneity of interventions and inconsistent inclusion of women with high risk may appear to limit the utility of the current evidence base. However, this is precisely the central contribution of our review: to highlight that, despite the volume of published trials, the current body of research lacks coherence, consistency in outcome reporting, and a focus on women most at risk for adverse maternal outcomes. This fragmentation has implications for clinical practice and research, particularly for populations with intersecting risk factors. We argue that this finding reveals an urgent need for more inclusive, systematically designed studies that address overlapping risk factors, consistently report maternal outcomes beyond GWG, and follow participants into the postpartum period. While overweight or obese BMI may increase risk for some pregnancies, over half of pregnant women in the US fall into these categories,^63–65^ yet not all of them experience APOs. This suggests that BMI alone is an imprecise and overly broad target for intervention. Moreover, women with higher BMI are more likely to gain excess gestational weight,^66^ which can contribute to PPWR and future CVD risk.^67, 68^ However, targeting BMI or GWG in isolation, especially in trials that exclude women with existing hypertension or diabetes, does not adequately address the broader drivers of maternal morbidity and mortality. While some studies in this review showed improvements in specific outcomes like GDM, HDPs or lipid profiles,^22, 38, 52, 53^ the overall evidence remains insufficient to conclude that GWG-focused interventions alone can improve comprehensive maternal health outcomes in the US, particularly among those at highest risk for APOs.

<u>Despite HDPs being the most dangerous type of APO^1^</u>, <u>only three studies in our review specifically recruited women with a history of HDPs or a prior hypertensive disorder.</u>^54, 59, 60^ This is notable given that the prevalence of HDPs and co-occurring GDM and HDPs is rising in the US, and these conditions are strong indicators of elevated CVD risk during pregnancy, postpartum, and into later life.^69–74^ HDPs specifically are responsible for a disproportionate share of maternal and fetal morbidity and mortality.^1, 75^ However, only 40% of the included studies in this review addressed risk factors other than obesity,^23–26, 29, 41, 44–46, 49, 53, 54, 56–60^ revealing a critical gap. This narrow focus limits our understanding of how to improve outcomes for women at the highest risk of APOs and future CVD.

Interestingly, the five studies added during our last literature search on June 30, 2024, four focused on either GDM or HDP risk, and none identified GWG as a primary outcome.^56–60^ Three of these newer studies implemented interventions beyond diet or physical activity, focusing on sleep,^58^ reducing sedentary behavior,^60^ and mindfulness training.^59^ This may signal an important shift toward a broader understanding of the behavioral and physiological contributors to pregnancy complications.

Studies combining diet and physical activity were generally more effective than those that focused on physical activity alone. In fact, the least effective interventions addressed only physical activity. Interventions that included dietary changes were more likely to impact health outcomes: three trials that incorporated nutrition components reported reduced HDP risk.^20, 52, 53^ For instance, one trial provided women with overweight or obesity a calorie-controlled balanced nutritional plan (18-24 kcal/kg/day; 40% carbohydrates, 30% protein, and 30% fat) along with nutritional counseling and food intake tracking.^52^ Another included formal nutritional counseling and diet therapy aligned with American Diabetes Association guidelines to reduce obstetrical complications in pregnant women with mild GDM.^53, 76^ A third included individualized calorie goals, healthy food education, increased physical activity, and self-weighing.^20^ While interventions that combined diet and physical activity were most likely to reduce GWG, most failed to improve other maternal outcomes. Critical measures like blood pressure, other biomarkers, or cesarean rates were often either not reported or unaffected. One study compared stretching to walking to prevent HDPs among pregnant women with prior HDPs,^54^ but while it observed some differences in transferrin levels and rates of preeclampsia vs. gestational hypertension, these results lacked clinical significance. Furthermore, this trial excluded women who developed chronic hypertension following preeclampsia, further narrowing its applicability.^54^

Although the primary focus of this review is on preventing APOs during pregnancy, several studies also extended their observations into the postpartum period, offering important insights into future work. Three interventions continued into the postpartum phase,^26, 44, 49^ while seven studies conducted follow-up assessments ranging from two weeks to one year postpartum.^32, 35, 47, 48, 50, 52, 55^ Some prenatal interventions, particularly those involving dietary changes^32^ or combined approaches with diet and physical activity,^26, 48^ were associated with reduced PPWR. One study^52^ also examined CVD-related postpartum outcomes, such as postpartum hypertension or preeclampsia, readmission to the hospital for postpartum CV-related complications (e.g., bleeding), providing a rare look into how prenatal behavioral interventions might influence maternal health beyond delivery. Recent research suggests that postpartum care may need to continue for at least a year postpartum or until the next pregnancy to address ongoing health risks, particularly for women who experienced an APO.^77, 78^ Future work should explore how extending behavioral interventions or follow-up into the postpartum period might improve long-term maternal outcomes, especially in populations at heightened risk for CVD.

<u>Potential reasons for null findings</u> include the unique challenges pregnancy presents, which may limit the effectiveness of lifestyle interventions that otherwise blood pressure, improve blood sugar, insulin levels, and body weight in non-pregnant populations.^7–10, 14^ Physical changes and discomfort during pregnancy often lead to altered eating patterns and reduced physical activity. Additionally, the mental strain associated with preparing for birth and caring for a newborn, compounded by physiological changes such as hormonal and metabolic shifts, may cause pregnant women to respond differently to behavioral interventions compared to non-pregnant people. To address this, interventions may need to be tailored specifically for pregnant populations by incorporating strategies that directly acknowledge and support mental health challenges during pregnancy. For example, integrating stress management techniques, emotional support, and flexible, pregnancy-safe activity options could improve intervention effectiveness. Moreover, recognizing that mental strain is a constant factor during pregnancy suggests that behavioral programs must be adaptable and trauma-informed, offering ongoing support rather than a one-time intervention.

Beyond individual factors, systemic barriers such as weight bias, structural racism, and structural determinants of health likely counteract intervention benefits. Most studies included in this review did not measure these factors. Only eight studies specifically recruited women from historically marginalized communities,^20, 26, 33, 38, 40–42, 47^ but none directly examined the influence of structural racism or social determinants on APOs. Additionally, the ways that race and ethnicity were categorized varied widely. Few studies reported how these data were collected (e.g., via self-report), and several conflated race and ethnicity, for instance, treating “Hispanic” as a racial category or excluding it entirely. This practice obscures within-group differences and erases the experiences of racially diverse Hispanic populations, including Black Hispanic communities. Some studies reported participant race as, “Other,” without clarification, further limiting interpretability. Moreover, 28 studies used BMI as an inclusion criterion despite its known limitations in accurately classifying overweight and obesity in certain racial and ethnic groups, especially among Black women.^79, 80^ These issues highlight how implicit bias and inconsistent data collection practices may shape study design, interpretation, and generalizability. Future research must explore how healthy behaviors might mitigate the harmful effects of structural racism and social inequality, and how these same forces limit engagement with or benefit from lifestyle interventions during pregnancy.

### Prevention efforts to reduce APOs ideally begin before pregnancy, and care for women who experience APOs should continue postpartum to reduce lifetime CVD risk

Risk factor modification prior to pregnancy is especially critical given that hypertension and diabetes have contributed to the rise in maternal morbidity in the US over the past 30 years.^81^ Healthy behaviors such as weight loss, adherence to the Dietary Approaches to Stop Hypertension (DASH) or Mediterranean diets, sodium reduction, potassium supplementation, increased physical activity, and reduced alcohol intake can be implemented before, during, and after pregnancy.^82, 83^ Combining stress management strategies with lifestyle interventions may offer additional benefits for hypertension prevention and control.^84^ This is important because perceived stress influences hypertension risk factors, especially among women of lower socioeconomic status.^85, 86^ Importantly, structural determinants of health such as limited access to safe exercise spaces or affordable nutritious food can exacerbate APO risk factors.

Prenatal care is an opportunity to engage patients early in pregnancy and provide risk assessment, psychosocial, cultural, and educational support with the ultimate goal of improving pregnancy outcomes.^87^ Midwives play a critical role in promoting healthy lifestyle interventions during pregnancy due to increased levels of contact and continuity with patients and recognition as a trusted source of advice and support.^88^ Prenatal care can also encourage women to be proactive in seeking appropriate primary health care postpartum. Unfortunately, many women do not receive appropriate assistance from their obstetric healthcare team to transition to primary care.^89, 90^ This is a lost opportunity to continue keeping postpartum women engaged in their health and healthcare to reduce the burden of CVD in women in the US, particularly for those who experienced APOs.

To better improve maternal outcomes, interventions should move beyond targeting the individual pregnant woman to implementing healthy lifestyle programs at the community level and developing multisector partnerships to address structural determinants of health.^91^ Collaboration between midwives and other obstetric health care providers, primary care providers, and patients will be necessary to ensure ongoing care into and beyond the postpartum years, especially for pregnant women at high risk for APOs and future CVD.^92^ Maternity care providers can help to ensure that postpartum women continue to monitor their health by providing a warm handoff to primary care. It is also vital to empower postpartum women to focus on their own health care in addition to that of their infants.

Despite mixed results regarding the benefit of behavioral interventions during pregnancy, there is clearly an overall deficit of such studies, particularly as there is a consensus that improving diet quality, increasing exercise, and reducing stress are important for overall health. Future research should investigate the impacts of behavioral interventions for high-risk pregnant women on maternal outcomes and long-term APO-related outcomes – with care to consider structural racism and determinants of health in the intervention – to develop the evidence base for improving maternal morbidity and mortality through health behavior change.

This systematic review has several limitations. First, we excluded follow-up reports of primary trials, which may have contained additional relevant data, particularly concerning postpartum outcomes. Future research should include these reports to gain a more comprehensive view of long-term intervention effects. Second, our focus on RCTs excluded many community-based behavioral programs that may offer valuable insight into culturally tailored or context-specific interventions for high-risk pregnant populations. Third, there was substantial heterogeneity in the included interventions in terms of content, intensity, duration, and follow-up, which limited our ability to compare specific strategies across trials. This variation is a common challenge in reviews of behavioral interventions and underscores the need for standardized reporting to support synthesis and comparison.

## CONCLUSION

This review reinforces that while behavioral interventions show some promise, the existing body of evidence lacks cohesion and relevance for the populations most in need. Nearly three-quarters of included studies excluded women with pre-existing hypertension or diabetes—conditions that are increasingly common among women of reproductive age and independently elevate the risk for APOs. Despite the complexity of CVD risk, most interventions centered narrowly on diet, physical activity, or weight, while underexploring other important behavioral contributors like sleep and stress. Moreover, studies rarely assessed clinically meaningful outcomes beyond gestational weight gain, such as blood pressure, glucose, or lipid levels—limiting our ability to determine how these interventions impact maternal cardiovascular health.

Future research should broaden inclusion criteria, use standardized outcome measures that reflect the multidimensional nature of CVD risk, and evaluate interventions beyond weight-related outcomes. Updated clinical practice guidelines should incorporate the best available evidence on health behaviors while acknowledging current gaps. Only through more inclusive, rigorous, and holistic approaches can we generate meaningful improvements in maternal health.

## Acknowledgements

Funding for this work was provided by Hatch Project #CA-D-HCE-2582-H, Addressing Multifactorial Influences on Pregnancy Outcomes to Promote Health Equity (Simmons PI) and 5R01NR017659 (Simmons PI). Dr. Keeton is currently supported by the National Center for Advancing Translational Sciences, National Institutes of Health, through grant number UL1 TR001860 and linked award KL2 TR001859. The content is solely the responsibility of the authors and does not necessarily represent the official views of the NIH.

## Data availability

The dataset generated and analyzed for this study are included in the article and supplement. Further data requests can be made to the corresponding author.

## Author contributions

J.E.P. and L.A.S. developed the concept and study design for this systematic review. M.V.N. generated the search strategy and performed the initial searches. J.E.P, I.A.D., N.S., A.K, and M.D.M.W. screened and reviewed all articles, performed full text extraction and bias analysis. I.E., T.K.T., H.L.H., and L.A.S. made decisions in cases where the reviewers had a disagreement. J.E.P, I.E., T.K.T., H.L.H., V.K., and L.A.S. drafted the manuscript. J.E.P., L.S.S., V.K., and L.A.S. edited the manuscript for critical content. All authors reviewed the manuscript, provided feedback, and approved the final manuscript.

## Statements and Declarations

The authors have no relevant financial or non-financial interests to disclose.

