## Supplement for "A systematic review of behavioral interventions to improve maternal outcomes for women in the United States at high risk for adverse pregnancy outcomes"

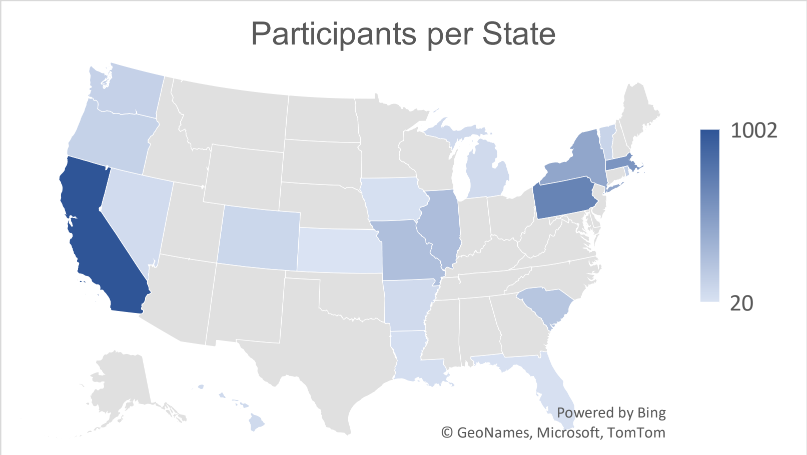


**Supplemental Figure S1.** Participants per state.

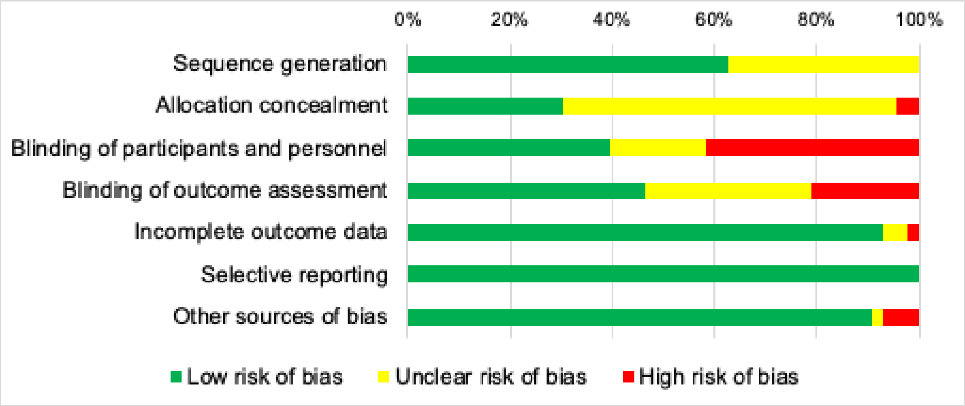


**Supplemental Figure S2.** Summary of risk of bias in included trials.

| | Sequence generation | Allocation concealment | Blinding of participants and personnel | Blinding of outcome assessment | Incomplete outcome data | Selective reporting | Other sources of bias |
|---|---|---|---|---|---|---|---|
| Allman 2021 | ? | ? | + | + | - | + | + |
| Basu 2021 | + | ? | + | + | + | + | + |
| Bublitz 2023 | + | ? | - | + | + | + | + |
| Cahill 2018 | + | + | + | + | + | + | + |
| Downs 2017 | + | ? | + | ? | + | + | + |
| Downs 2021 | + | - | - | ? | + | + | + |
| Durnwald 2016 | + | - | - | - | + | + | + |
| Ferrara 2011 | + | ? | + | ? | + | + | + |
| Ferrara 2020 | + | ? | + | + | + | + | + |
| Gallagher 2018 | ? | ? | + | + | + | + | + |
| Gibbs 2024 | + | ? | - | ? | + | + | + |
| Goletzke 2021 | ? | ? | ? | ? | + | + | + |
| Hawkins 2015 | ? | ? | - | + | + | + | + |
| Hernandez 2016 | ? | ? | - | - | ? | + | + |
| Hernandez 2023 | + | ? | - | + | + | + | ? |
| Herring 2016 | + | + | + | ? | + | + | + |
| Holmes 2020 | + | + | - | ? | + | + | + |
| Homko 2012 | ? | ? | - | - | + | + | + |
| Hull 2020 | + | ? | ? | ? | + | + | - |
| Jaworsky 2023 | ? | ? | - | + | + | + | + |
| Kong 2014 | + | ? | + | + | + | + | + |
| Landon 2009 | + | ? | ? | ? | + | + | + |
| LeBlanc 2021 | ? | ?/+ | + | ? | + | + | + |
| Liu 2021 | + | + | - | + | + | + | + |
| Mackeen 2022 | + | ? | + | + | + | + | + |
| Nobles 2015 | + | ? | + | + | + | + | + |
| Osmundson 2016 | + | + | - | - | + | + | + |
| Palnati 2021 | ? | ? | + | + | + | + | + |
| Pauley 2018 | ? | ? | ? | ? | + | + | + |
| Phelan 2018 | + | ? | - | + | + | + | + |
| Phillips 2019 | + | ? | - | - | + | + | + |
| Reader 2006 | ? | ? | ? | + | + | + | + |
| Redman 2017 | ? | + | + | - | + | + | + |
| Rhodes 2010 | ? | + | + | ? | + | + | - |
| Sacks 2006 | + | ? | ? | ? | + | + | - |
| Sugino 2022 | ? | ? | - | - | + | + | + |
| Thomas 2022 | ? | ? | + | + | + | + | + |
| Thornton 2009 | + | + | - | - | + | + | + |
| Trout 2016 | ? | ? | ? | ? | + | + | + |
| Twedt 2023 | + | + | - | - | ? | + | + |
| VanHorn 2018 | + | + | + | + | + | + | + |
| Vesco 2014 | + | + | - | + | + | + | + |
| Yeo 2008 | + | + | ? | + | + | + | + |

**Supplemental Figure S3.** Risk of bias for each included trial.

**Supplemental Figure S4.** PRISMA checklist – included as a pdf attachment in landscape view.

**Supplemental Table S1.** Details of all included trials – included as a pdf attachment in landscape view.

**Supplemental Table S2. Search strategy**

Ovid MEDLINE (via PubMed):

| Search | Query | Results 12.03.21 | Results 04.05.23 |
|---|---|---|---|
| #1 | ("Hypertension"[Mesh:NoExp] OR "Hypertension, Pregnancy-Induced"[Mesh] OR hypertension[tiab] OR hypertensive[tiab] OR "high blood pressure"[tiab] OR eclampsia[tiab] OR eclampsias[tiab] OR "HELLP Syndrome"[Mesh] OR "HELLP Syndrome"[tiab] OR "EPH Complex"[tiab] OR "EPH Gestosis"[tiab] OR "Toxemia"[Mesh] OR toxemia[tiab] OR toxemias[tiab] OR "pre eclampsia"[tiab] OR "preeclampsia"[tiab] OR "Pregnancy Complications, Cardiovascular"[Mesh] OR "Obstetric Labor, Premature"[Mesh] OR "pre-term labor"[tiab] OR "preterm labor"[tiab] OR "premature labor"[tiab] OR "premature obstetric labor"[tiab] OR "Heart Defects, Congenital"[Mesh] OR "congenital heart defect"[tiab] OR "congenital heart defects"[tiab] OR "congenital heart disease"[tiab] OR "congenital heart diseases"[tiab] OR "heart abnormality"[tiab] OR "heart abnormalities"[tiab] OR "Arrhythmias, Cardiac"[Mesh] OR "cardiac dysrhythmia"[tiab] OR arrhythmia[tiab] OR arrhythmias[tiab] OR arrythmia[tiab] OR arrythmias[tiab] OR "Heart Valve Diseases"[Mesh] OR "heart valve disease"[tiab] OR "heart valve diseases"[tiab] OR "valvular heart disease"[tiab] OR "valvular heart diseases"[tiab] OR "Cardiomyopathies"[Mesh] OR Cardiomyopathy[tiab] OR Cardiomyopathies[tiab] OR "Myocardial Disease"[tiab] OR "Myocardial Diseases"[tiab] OR Myocardiopathies[tiab] OR Myocardiopathy[tiab] OR "Hypertension, Pulmonary"[Mesh] OR "Pulmonary Hypertension"[tiab] OR "Coronary Artery Disease"[Mesh] OR "Coronary Artery Disease"[tiab] OR "Coronary Artery Diseases"[tiab] OR "Coronary Arteriosclerosis"[tiab] OR "Coronary Arterioscleroses"[tiab] OR "Coronary Atheroscleroses"[tiab] OR "Coronary Atherosclerosis"[tiab] OR "Heart Transplantation"[Mesh] OR "Heart Grafting"[tiab] OR "Heart Graftings"[tiab] OR "Heart Transplant"[tiab] OR "Heart Transplants"[tiab] OR "Heart Transplantation"[tiab] OR "Heart Transplantations"[tiab] OR "Cardiac Transplantation"[tiab] OR "Cardiac Transplantations"[tiab] OR "Obesity"[Mesh:NoExp] OR "Obesity, Maternal"[Mesh] OR "obesity"[tiab] OR "obese"[tiab] OR "gestational weight gain"[tiab] OR "Growth Disorders"[Mesh] OR "growth disorder"[tiab] OR "Fetal Growth Retardation"[Mesh] OR "Intrauterine Growth Retardation"[tiab] OR "Intrauterine Growth Restriction"[tiab] OR "Fetal Growth Restriction"[tiab] OR "Diabetes Mellitus"[Mesh:NoExp] OR "Diabetes Mellitus, Type 2"[Mesh] OR "Diabetes, Gestational"[Mesh] OR diabetes[tiab] OR diabetic[tiab] OR "Polycystic Ovary Syndrome"[Mesh] OR "Stein | 179,069 | 193,886 |

| | | | |
|---|---|---|---|
| | Leventhal Syndrome"[tiab] OR "Sclerocystic Ovarian Degeneration"[tiab] OR "Sclerocystic Ovary Syndrome"[tiab] OR "Polycystic Ovarian Syndrome"[tiab] OR "Polycystic Ovary Syndrome"[tiab] OR "Sclerocystic Ovaries"[tiab] OR "Sclerocystic Ovary"[tiab] OR "Hypothyroidism"[Mesh] OR "Hypothyroidism"[tiab] OR "Hypothyroidisms"[tiab] OR "Thyroid Stimulating Hormone Deficiency"[tiab] OR "Thyroid Stimulating Hormone Deficiencies"[tiab] OR "TSH Deficiency"[tiab] OR "TSH Deficiencies"[tiab] OR "Hashimoto's Thyroiditis"[tiab] OR "Hashimotos Thyroiditis"[tiab] OR "Hashimoto Thyroiditis"[tiab] OR "Hashimoto's Thyroiditides"[tiab] OR "Hashimotos Thyroiditides"[tiab] OR "Hashimoto Thyroiditides"[tiab] OR "Hashimoto's Syndrome"[tiab] OR "Hashimotos Syndrome"[tiab] OR "Hashimoto Syndrome"[tiab] OR "Hashimoto's Struma"[tiab] OR "Hashimotos Struma"[tiab] OR "Hashimoto Struma"[tiab] OR "Hashimoto's Disease"[tiab] OR "Hashimotos Disease"[tiab] OR "Hashimoto Disease"[tiab] OR "Chronic Lymphocytic Thyroiditis"[tiab] OR "Chronic Lymphocytic Thyroiditides"[tiab]) AND ("Pregnancy"[Mesh:NoExp] OR "Pregnant Women"[Mesh] OR "Pregnancy, High-Risk"[Mesh] OR pregnancy[ti] OR pregnancy[ot] OR pregnant[ti] OR pregnant[ot] OR pregnancies[ti] OR pregnancies[ot] OR maternal[ti] OR maternal[ot] OR "Reproductive History"[Mesh] OR parous[tiab] OR parity[tiab] OR gravidities[tiab] OR multiparity[tiab] OR nulligravidity[tiab] OR primigravidity[tiab] OR multigravidity[tiab]) | | |
| #2 | "weight management"[tw] OR "stress management"[tw] OR "physical activity"[tw] OR exercise[tw] OR "diet"[tw] OR "mindfulness"[tw] OR "quiet rest"[tw] OR "music therapy"[tw] OR "aromatherapy"[tw] OR "relaxation therapy"[tw] OR "acupuncture"[tw] OR "acupressure"[tw] OR "massage"[tw] OR "device-guided slow breathing"[tw] OR "hypnosis"[tw] OR "yoga"[tw] | 996,960 | 1,078,830 |
| #3 | #1 AND #2 | 11,011 | 12,230 |
| #4 | #3 AND ("controlled clinical trial"[pt] OR "randomized controlled trial"[pt] OR "randomized"[tiab] OR "randomised"[tiab] OR "randomization"[tiab] OR "randomisation"[tiab] OR "randomly"[tiab] OR placebo*[tiab] OR "Clinical Trials as Topic"[mh] OR trial[ti] OR "Clinical Trial, Phase III"[pt] OR "Clinical Trials, Phase III as Topic"[mh]) NOT (Editorial[pt] OR Letter[pt] OR Case Reports[pt] OR Comment[pt]) NOT (animals[mh] NOT humans[mh]) | 1,537 | 1,733 |
| #5 | #4 AND English[lang] | 1,480 | 1,676 |

Cochrane Central Register of Controlled Trials:

| **Search** | **Query** | **Results 12/03/2021** | **Results 04/05/2023** |
|---|---|---|---|

| | | | |
|---|---|---|---|
| #1 | ((hypertension OR hypertensive OR high blood pressure OR eclampsia OR eclampsias OR Hemolysis Elevated Liver Enzymes Lowered Platelets OR HELLP Syndrome OR EPH Complex OR EPH Gestosis OR toxemia OR toxemias OR pre eclampsia OR preeclampsia OR pre-term labor OR preterm labor OR premature labor OR premature obstetric labor OR congenital heart defect OR congenital heart defects OR congenital heart disease OR congenital heart diseases OR heart abnormality OR heart abnormalities OR cardiac dysrhythmia OR arrhythmia OR arrhythmias OR arrythmia OR arrythmias OR heart valve disease OR heart valve diseases OR valvular heart disease OR valvular heart diseases OR Cardiomyopathy OR Cardiomyopathies OR Myocardial Disease OR Myocardial Diseases OR Myocardiopathies OR Myocardiopathy OR Pulmonary Hypertension OR Coronary Artery Disease OR Coronary Artery Diseases OR Coronary Arteriosclerosis OR Coronary Arterioscleroses OR Coronary Atheroscleroses OR Coronary Atherosclerosis OR Heart Grafting OR Heart Graftings OR Heart Transplant OR Heart Transplants OR Heart Transplantation OR Heart Transplantations OR Cardiac Transplantation OR Cardiac Transplantations OR obesity OR obese OR gestational weight gain OR growth disorder OR Intrauterine Growth Retardation OR Intrauterine Growth Restriction OR Fetal Growth Restriction OR diabetes OR diabetic OR Stein Leventhal Syndrome OR Sclerocystic Ovarian Degeneration OR Sclerocystic Ovary Syndrome OR Polycystic Ovarian Syndrome OR Polycystic Ovary Syndrome OR Sclerocystic Ovaries OR Sclerocystic Ovary OR Hypothyroidism OR Hypothyroidisms OR Thyroid Stimulating Hormone Deficiency OR Thyroid Stimulating Hormone Deficiencies OR TSH Deficiency OR TSH Deficiencies OR Hashimoto's Thyroiditis OR Hashimotos Thyroiditis OR Hashimoto Thyroiditis OR Hashimoto's Thyroiditides OR Hashimotos Thyroiditides OR Hashimoto Thyroiditides OR Hashimoto's Syndrome OR Hashimotos Syndrome OR Hashimoto Syndrome OR Hashimoto's Struma OR Hashimotos Struma OR Hashimoto Struma OR Hashimoto's Disease OR Hashimotos Disease OR Hashimoto Disease OR Chronic Lymphocytic Thyroiditis OR Chronic Lymphocytic Thyroiditides) AND (women OR woman* OR female OR females) AND (pregnancy OR pregnant OR pregnancies OR parous OR parity OR gravidities OR multiparity OR | 15,434 | 17,494 |

| | | | |
|---|---|---|---|
| | nulligravidity OR nulligravidities OR primigravidity OR primigravidities OR multigravidity OR multigravidities)):ti,ab,kw<br>*(Word variations have been searched)* | | |
| #2 | (weight management OR stress management OR physical activity OR exercise OR diet OR mindfulness OR quiet rest OR music therapy OR aromatherapy OR relaxation therapy OR acupuncture OR acupressure OR massage OR device-guided slow breathing OR hypnosis OR yoga):ti,ab,kw<br>*(Word variations have been searched)* | 251,859 | 284,522 |
| #3 | #1 AND #2<br>*(in Trials)*<br>*Language: English* | 3,773<br><br>**Source:**<br>Embase: 1,615<br>PubMed: 1,337<br>ICTRP: 749<br>CT.gov: 666<br>CINAHL: 32 | 4,253<br><br>**Source:**<br>Embase: 1,817<br>PubMed: 1,477<br>ICTRP: 905<br>CT.gov: 758<br>CINAHL: 53 |

Ovid MEDLINE (via PubMed):

| **Search** | **Query** | **Results 5.1.24** |
|---|---|---|
| #1 | (((((((diabetes) OR (hypertension)) OR (preeclampsia)) OR (adverse pregnancy outcomes)) OR (physical activity)) OR (diet)) OR (mindfulness) AND ((randomizedcontrolledtrial[Filter]) AND (2023/1/1:2024/1/1[pdat]))) AND (pregnancy) Filters: Randomized Controlled Trial, from 2023/1/1 - 2024/6/30 | 452 |
