## Supplemental Table S1 for "A systematic review of behavioral interventions to improve maternal outcomes for women in the United States at high risk for adverse pregnancy outcomes"

| First author, year of publication, (citation) | Title | Aim of Study | Number of participants | Length of intervention | When intervention ended | When monitoring participants ended | High risk feature trial was designed for | Intervention type | Outcome of intervention | Method of Recruitment | Race and ethnicity reported as separate categories | Notes on race and ethnicity reporting | Excluded women with pre-existing CVD factors other than feature trial was designed for | BMI requirement |
|---|---|---|---|---|---|---|---|---|---|---|---|---|---|---|
| Allman 2021 | Markers of Branched-Chain Amino Acid Catabolism are Not Affected by Exercise Training in Pregnant Women with Obesity | Determine the effect of a moderate-intensity exercise intervention during pregnancy on maternal circulating branched-chain amino acids (BCAAs) and markers of BCAA catabolism, and their associations with insulin resistance. | 80 | ~24 weeks | prenatal | prenatal | Overweight/obesity | Physical activity | Physical activity was safe for pregnant participants (intervention feasible) | Clinic | Yes | Race: Caucasian, African American, More than One/Missing | yes | obese |
| Basu 2021 | Dietary Blueberry and Soluble Fiber Supplementation Reduces Risk of Gestational Diabetes in Women with Obesity in a Randomized Controlled Trial | Determine the effect of blueberry and soluble fiber supplementation on GDM risk and GWG for pregnant people with obesity | 34 | ~18 weeks | prenatal | prenatal | Overweight/obesity; Gestational diabetes | Diet | Reduced GWG; lower diastolic blood pressure | Clinic | No | Race: African American and Hispanic | yes | obese |
| Bublitz 2023 | Feasibility, acceptability, and preliminary effects of mindfulness training on antenatal blood pressure | Test the feasibility, acceptability, and effects of minfulness training on rates of HDP among pregnant women at risk for HDP | 29 | 8 weeks | prenatal | prenatal | Hypertensive disorders of pregnancy | Other: mindfulness training | Feasibility; improved blood pressure | Clinic | No | Only provided White race and Hispanic ethnicity | no | none |
| Cahill 2018 | Weight Control Program and Gestational Weight Gain in Disadvantaged Women with Overweight or Obesity: a Randomized Clinical Trial | Evaluate whether an optimized, home-based, lifestyle weight management program could affect GWG and improve maternal and neonatal outcomes in socioeconomically disadvantaged African American pregnant people with overweight or obesity at the beginning of pregnancy. | 267 | 20 weeks (10 biweekly home visits during pregnancy) | prenatal | prenatal | Overweight/obesity | Diet; Physical activity | Reduced GWG | Clinic | Yes | Only Black/African American race reported since this was conducted in this population | yes | overweight or obese |
| Downs 2017 | Randomized face-to-face vs. home exercise interventions in pregnant women with gestational diabetes | Evaluate the effects of a semi-intensive, face-to-face exercise intervention and a minimum-contact, home-based exercise intervention compared to the standard of care (control) on exercise behavior, its motivational determinants, blood glucose levels, and insulin use of women with GDM. | 41 | 12 weeks | prenatal | prenatal | Gestational diabetes | Diet; Physical activity | Reduced blood glucose levels | Clinic | No | Race and ethnicity: non-Hispanic White, Asian American, Hispanic, American Indian, Other | no | none |
| Downs 2021 | Adaptive, behavioral intervention impact on weight gain, physical activity, energy intake, and motivational determinants: results of a feasibility trial in pregnant women with overweight/obesity | Reduce GWG in pregnant people with overweight/obesity through mHealth targeted to diet and physical activity | 31 | 8-12 weeks through 36 weeks | prenatal | prenatal | Overweight/obesity | Diet; Physical activity | Intervention feasible | Social media or online | No | Race: Caucasian/white , Asian | yes | overweight or obese |
| Durnwald 2016 | A Randomized Clinical Trial of an Intensive Behavior Education Program in Gestational Diabetes Mellitus Women Designed to Improve Glucose Levels on the 2-Hour Oral Glucose Tolerance Test | Evaluate whether pregnant people with GDM enrolled in an intensive behavior educational program demonstrate lower mean fasting glucose levels on the 2-hour 75g oral glucose tolerance test at 6-12 weeks postpartum compared to routine GDM management | 101 | 30 weeks through 6-12 weeks postpartum | postpartum | postpartum | Gestational diabetes | Diet; Physical activity | Null | Clinic | Yes | Reported correctly | yes | none |
| Ferrara 2011 | A pregnancy and postpartum lifestyle intervention in women with gestational diabetes mellitus reduces diabetes risk factors: a feasibility randomized control trial | Evaluate feasibility of a lifestyle intervention initiated soon after GDM diagnosis and continuing postpartum. The intervention goals were 1) help postpartum people return to prepregnancy weight or 5% below for postpartum people with overweight/obesity | 197 | GDM diagnosis through 1 year postpartum | postpartum | postpartum | Gestational diabetes | Diet; Physical activity | Decreased dietary fat intake | Clinic | No | Reported correctly | yes | none |
| Ferrara 2020 | A telehealth lifestyle intervention to reduce excess gestational weight gain in pregnant women with overweight or obesity (GLOW): a randomised, parallel-group, controlled trial | Reduce excess GWG through a behavioral lifestyle intervention adapted from the Diabetes Prevention Program | 394 | 8-15 weeks through 29-38 weeks | prenatal | prenatal | Overweight/obesity | Diet; Physical activity; Other: stress management | Reduced GWG | Clinic | No | Race and ethnicity: Asian, White, Hispanic, African American, Multiracial/Other | yes | overweight or obese |

| | | | | | | | | | | | | | | |
|---|---|---|---|---|---|---|---|---|---|---|---|---|---|---|
| Gallagher 2018 | Greater Neonatal Fat-Free Mass and Similar Fat Mass Following a Randomized Trial to Control Excess Gestational Weight Gain | Determine the effectiveness of controlling GWG in the second and third trimesters on neonate body composition | 210 | 15 weeks through delivery | prenatal | prenatal | Overweight/obesity | Diet; Physical activity; Other: counseling/coaching | Reduced GWG | Clinic | Yes | Race: White, Black, Other, More than One, Unknown | no | overweight or obese |
| Gibbs 2024 | The sedentary behavior reduction in pregnancy intervention (SPRING) pilot and feasibility randomized trial | To demonstrate feasibility, acceptability, and initial efficacy of a lower intensity intervention targeting reduced sedentary behavior and increased standing and steps | 51 | Clinic and online | prenatal | prenatal | High risk for any APO, high sedentary behavior | Physical activity and Other: counseling/coaching | Reduced sedentary time | Clinic and online | Yes | Overall hispanic yes/no, but no ethnicity data provided for individual race data | yes | none |
| Goletzke 2021 | Effect of a Low-Glycemic Load Diet Intervention on Maternal and Pregnancy Outcomes in Obese Pregnant Women | Compare effects of low vs high glycemic diets during the second half of pregnancy on maternal glucose metabolism and pregnancy outcomes in women with overweight or obesity | 109 | 20 weeks through delivery | prenatal | prenatal | Overweight/obesity | Diet | Reduced GWG | Clinic | No | Listed all as ethnicities: Hispanic, African American, Caucasian, Asian, Mixed Ethnicity, Hawaiian/Pacific Islander | yes | obese |
| Hawkins 2015 | A pregnancy lifestyle intervention to prevent gestational diabetes risk factors in overweight Hispanic women: a feasibility randomized controlled trial | Evaluate the feasibility of a lifestyle intervention among Hispanic pregnant people with overweight or obesity who have a higher risk of GDM. The aims of the intervention were to reduce excess gestational weight gain, increase postpartum weight loss and improve maternal metabolic status in this population. | 68 | ~26 weeks | prenatal | postpartum | Overweight/obesity | Diet; Physical activity | Increased minutes of vigorous physical activity | Clinic | Yes | 100% Hispanic, unsure if authors asked about identifying with other racial categories | yes | overweight or obese |
| Hernandez 2016 | Women With Gestational Diabetes Mellitus Randomized to a Higher-Complex Carbohydrate/Low-Fat Diet Manifest Lower Adipose Tissue Insulin Resistance, Inflammation, Glucose, and Free Fatty Acids: a Pilot Study | Determine if a high-complex carb/lower fat diet improves maternal insulin resistance, adipose tissue lipolysis and infant adiposity | 12 | 30-31 weeks until delivery | prenatal | prenatal | Overweight/obesity; Gestational diabetes | Diet | Reduced fasting glucose; reduced free fatty acids | Clinic | No | Race: White, Asian, Hispanic | yes | overweight or obese |
| Hernandez 2023 | Randomization to a Provided Higher-Complex-Carbohydrate Versus Conventional Diet in Gestational Diabetes Mellitus Results in Similar Newborn Adiposity | Determine if following a CHOICE diet compared to a conventional diet would be associated with improved insulin resistance and 24-hr gylcemia in pregnant participants with GDM | 46 | 30-31 weeks through delivery | prenatal | prenatal | Overweight/obesity; Gestational diabetes | Diet; Physical activity | Null | Clinic | No | Overall hispanic yes/no, but no ethnicity data provided for individual race data | yes | overweight or obese |
| Herring 2016 | Preventing excessive gestational weight gain among African American women: a randomized clinical trial | Determine whether a technology-based behavioral weight control intervention would be effective among low-income African American pregnant people with overweight or obesity for decreasing excess GWG | 66 | 20 weeks through delivery | prenatal | prenatal | Overweight/obesity | Diet; Physical activity; Other: self-weighing | Reduced GWG | Clinic | Yes | Only recruited self-identified African-American participants, unsure if authors asked about identifying with Hispanic ethnicity | no | overweight or obese |
| Holmes 2020 | Effect of a short message service intervention on excessive gestational weight gain in a low-income population: a randomized controlled trial | Assess the effectiveness of an 18-week SMS intervention promoting nutrition and physical activity delivered to a low-income population of predominantly pregnant people with overweight/obese in Hawaii on reducing excessive GWG | 83 | 18 weeks | prenatal | prenatal | Overweight/obesity | Diet; Physical activity | Null | Clinic | No | Race and ethnicity: Asian, American Indian, Black, Hispanic, Native Hawaiian, Pacific Islander, White and they note that 45/82 (54.2%) of participants identified with multiple races and ethnicities | no | 20-45 |
| Homko 2012 | Impact of a telemedicine system with automated reminders on outcomes in women with gestational diabetes mellitus | Examine the impact of an enhanced telemedicine system on glucose control and pregnancy outcomes in pregnant people with GDM | 80 | GDM diagnosis through delivery (no later than 33 weeks for enrollment) | prenatal | prenatal | Gestational diabetes | Other: diabetes self management (self-monitoring glucose) | Contact between participants with GDM and their providers increased (intervention feasible) | Clinic | No | Race and Ethnicity: White, African American, Latino/Hispanic, Asian and Other | yes | none |

| | | | | | | | | | | | | | | |
|---|---|---|---|---|---|---|---|---|---|---|---|---|---|---|
| Hull 2020 | The effect of high dietary fiber intake on gestational weight gain, fat accrual, and postpartum weight retention: a randomized clinical trial | Assess effectiveness of a single goal high fiber diet intervention to prevent excessive GWG compared to usual care | 20 | 12 weeks | prenatal | postpartum | Overweight/obesity | Diet | Reduced GWG; reduced PPWR | Clinic | No | Only 2 categories, Hispanic/Latino and non-Hispanic/Latino | yes | ≥22 |
| Jaworsky 2023 | Effects of an Eating Pattern Including Colorful Fruits and Vegetables on Management of Gestational Diabetes: A Randomized Controlled Trial | Determine if increasing the consumption of berries and green leafy vegetables, as well as increasing postprandial physical activity, would improve cardiometabolic profiles in pregnant women with GDM. | 38 | 12 weeks | prenatal | prenatal | Gestational diabetes | Diet; Physical activity | Improved blood glucose, decreased serum IL-6, improved HDL cholesterol | Clinic | No | Only ethnicity provided (100% Hispanic) | no | none |
| Kong 2014 | A Pilot Walking Program Promotes Moderate-Intensity Physical Activity during Pregnancy | Determine if a walking intervention in pregnancy reduced PPWR in pregnant people with overweight/obesity. Additionally, determine if previously nonexercising pregnant people with overweight or obesity could increase moderate intensity PA participation via a walking intervention and if this intervention improved pregnancy and birth outcomes | 47 | 15-35 weeks | prenatal | postpartum | Overweight/obesity | Physical activity | Increased minutes of walking | Social media or online | No | Only White reported | yes | overweight or obese |
| Landon 2009 | A multicenter, randomized trial of treatment for mild gestational diabetes | Determine if treatment of mild GDM reduces perinatal and obstetrical complications | 958 | ~24 weeks through delivery | prenatal | prenatal | Gestational diabetes | Diet; Other: self-monitoring blood glucose | Reduced GWG; lower rates of HDP; GDM controlled | Clinic | No | Black, White, Asian, Hispanic, Other | yes | none |
| LeBlanc 2021 | Weight loss prior to pregnancy and subsequent gestational weight gain: Prepare, a randomized clinical trial | Determine whether prepregnancy weight loss reduces GWG and improves pregnancy outcomes | 169 | prepregnancy (up to 2 years) through delivery | prenatal | prenatal | Overweight/obesity | Diet; Physical activity | Null | Social media or online | Yes | Race Categories: White, Asian, Black, More than One, Did not Report | no | ≥27 |
| Liu 2021 | A Behavioral Lifestyle Intervention to Limit Gestational Weight Gain in Pregnant Women with Overweight and Obesity | Examine the impact of a pregnancy and postpartum behavioral lifestyle intervention (vs. standard care) on postpartum weight retention during the first year after delivery among white and African American women with overweight or obesity. | 219 | ≤18 weeks through delivery | prenatal | prenatal | Overweight/obesity | Diet; Physical activity; self-monitoring | Reduced rates of HDP | Social media or online | No | White, Black/African American (recruited to reflect South Carolina's population) | no | overweight or obese |
| Mackeen 2022 | Encouraging appropriate gestational weight gain in high-risk gravida: a randomized controlled trial | Decrease excessive GWG among women with pregestational obesity | 224 | <17 weeks through delivery | prenatal | prenatal | Overweight/obesity | Diet; Physical activity | Reduced GWG for participants with class III obesity | Clinic | No | 91.8% intervention and 86% control are non-Hispanic white. Other races and ethnicities not reported | no | obese |
| Nobles 2015 | Effect of an exercise intervention on gestational diabetes mellitus: a randomized controlled trial | Examine the effect of a prenatal exercise intervention on GDM | 241 | 12 weeks | prenatal | prenatal | Overweight/obesity; Gestational diabetes | Physical activity | Null | Clinic | No | Only Hispanic and Non-Hispanic categories reported | yes | overweight or obese |
| Osmundson 2016 | Early Screening and Treatment of Women with Prediabetes: a Randomized Controlled Trial | Examine whether treatment of pregnant people with a first-trimester A1C of 5.7 to 6.4% reduces the incidence of GDM compared with usual prenatal care. | 83 | first trimester through delivery | prenatal | prenatal | Gestational diabetes | Diet; Other: Self-monitoring | Reduced GDM rates for participants without obesity; lower A1c levels | Clinic | No | Race: White, Black, Hispanic, Asian | yes | none |
| Palnati 2021 | The Impact of a Lifestyle Intervention on Postpartum Weight Retention Among At-Risk Hispanic Women | Examine the effect of a lifestyle intervention on postpartum weight retention in Hispanic pregnant people with abnormal glucose tolerance in their current pregnancy. | 204 | 29 weeks through 1 year postpartum | postpartum | postpartum | Gestational diabetes | Diet; Physical activity | Reduced PPWR | Clinic | Yes | 100% Hispanic participants "Hispanic ethnicity was identified via self-report in the manner of the U.S. Census." | yes | none |
| Pauley 2018 | Gestational Weight Gain Intervention Impacts Determinants of Healthy Eating and Exercise in Overweight/Obese Pregnant Women | Establish the feasibility of the intervention components and the extent to which the theory of planned behavior and self-regulation constructs motivated pregnant people with overweight/obesity to engage in exercise and healthy eating behaviors. | 17 | 6 week intervention | prenatal | prenatal | Overweight/obesity | Diet; Physical activity | Improved attitudes and self-regulation regarding exercise and nutritious eating | Clinic | No | Ethnicity (Hispanic/non-Hispanic) is separted, but Caucasian is the only race category reported | unknown | overweight or obese |

| | | | | | | | | | | | | | | |
|---|---|---|---|---|---|---|---|---|---|---|---|---|---|---|
| Phelan 2018 | Randomized controlled clinical trial of behavioral lifestyle intervention with partial meal replacement to reduce excessive gestational weight gain | Determine the efficacy of a multicomponent behavioral lifestyle intervention with partial meal replacement on GWG rates in Hispanic and non-Hispanic pregnant people with overweight or obesity | 257 | 9-16 weeks through delivery | prenatal | prenatal | Overweight/obesity | Diet; Physical activity | Reduced GWG; lower blood trigylceride levels | Clinic | Yes | Ethnicity (Hispanic/non-Hispanic) is separated, races are American Indian, Alaskan Native, Asian, Black or African American, Native Hawaiian or Pacific Islander, White, and Other. Participants could select multiple races/ethnicities | yes | overweight or obese |
| Phillips 2019 | Combined financial incentives and behavioral weight management to enhance adherence with gestational weight gain guidelines: a randomized controlled trial | Determine the combined efficacy of financial incentives and behavioral weight management on adherence to GWG guidelines in pregnant people with overweight or obesity | 124 | 16 weeks through delivery | prenatal | postpartum | Overweight/obesity | Diet; Other: counseling/coaching | Null | Clinic | No | White is only category reported | no | overweight or obese |
| Reader 2006 | Impact of gestational diabetes mellitus nutrition practice guidelines implemented by registered dietitians on pregnancy outcomes | Determine if nutrition care delivered by RDs using nutrition practice guidelines for GDM results in different care and better pregnancy outcomes compared with usual nutrition care provided by RDs | 215 | GDM diagnosis through delivery (approximately 9 weeks) | prenatal | prenatal | Gestational diabetes | Diet | Less insulin required for GDM treatment | Clinic | No | Ethnicity: White, Hispanic, African American, Other | yes | none |
| Redman 2017 | Effectiveness of SmartMoms, a Novel eHealth Intervention for Management of Gestational Weight Gain: Randomized Controlled Pilot Trial | Decrease the proportion of pregnant people who exceed the IOM GWG guidelines | 54 | 13 wks 5 days through delivery | prenatal | postpartum | Overweight/obesity | Diet; Physical activity | Reduced GWG | Clinic | No | Black, White, and Other are the only categories reported. No ethnicity data reported | yes | overweight or obese |
| Rhodes 2010 | Effects of a low-glycemic load diet in overweight and obese pregnant women: a pilot randomized controlled trial | Examine the effects of a low glycemic load diet in overweight and obese pregnant people with the provision of specific foods to enhance treatment fidelity. | 46 | 13-28 weeks through delivery | prenatal | prenatal | Overweight/obesity | Diet | Reduced blood levels of triglycerides, total cholesterol and C-reactive protein | Clinic | No | Race: non-Hispanic white, non-Hispanic Black, Hispanic, Asian, More than one, Missing, Other | yes | overweight or obese |
| Sacks 2006 | Managing type I diabetes in pregnancy: how near normal is necessary? | This was a feasibility study to determine if pregnant people with type I diabetes managed with liberal target glucose values will have a decreased frequency of hypoglycemia with no differences in adverse outcomes compared with tightly controlled subjects. | 22 | <13 weeks gestation through delivery | prenatal | prenatal | Type 1 diabetes | Diet; Other: glucose self-monitoring, counseling/coaching | Improved glucose levels; reduced hypoglycemia | Clinic | No | Race: Caucasian, Other | unknown | none |
| Sugino 2022 | A maternal higher-complex carbohydrate diet increases bifidobacteria and alters early life acquisition of the infant microbiome in women with gestational diabetes mellitus | Determine if consumption of a higher complex carbohydrate/lower fat diet would favorably alter maternal and infant microbiome composition compared to a conventional diet in pregnant people with GDM | 34 | 30-31 weeks through delivery | prenatal | prenatal | Overweight/obesity; Gestational diabetes | Diet | Improved maternal gut microbiome | Clinic | No | Caucasian, Asian, Native American or Alaska Indian, Other | yes | overweight or obese |
| Thomas 2022 | A Web-Based mHealth Intervention With Telephone Support to Increase Physical Activity Among Pregnant Patients With Overweight or Obesity: Feasibility Randomized Controlled Trial | Test the acceptability and feasibility of a pilot mHealth lifestyle intervention for pregnant patients with overweight or obesity to promote moderate intense physical activity, encourage guideline-concordant GWG, and inform the design of a larger pragmatic cluster randomized controlled trial. | 68 | 8-15 weeks through delivery | prenatal | prenatal | Overweight/obesity | Diet; Physical activity | Improved self monitoring of weight; increased physical activity levels | Clinic | Yes | Reported correctly | yes | overweight or obese |

| | | | | | | | | | | | | | | |
|---|---|---|---|---|---|---|---|---|---|---|---|---|---|---|
| Thornton 2009 | Perinatal outcomes in nutritionally monitored obese pregnant women: a randomized clinical trial | (1) Compare perinatal outcomes of pregnant people with obesity treated in the conventional manner compared to nutritional monitoring; (2) Determine the effects of weight stabilization in pregnant people with obesity on perinatal morbidity and birth weight of newborns; (3) Determine perinatal differences in the study group's adherence vs non-adherence to a prescribed nutritional regimen applicable to the general practice of obstetrics; (4) Evaluate perinatal outcomes of pregnant people with obesity who had gained 15 pounds or more during their pregnancy compared to those who gained fewer than 15 pounds, irrespective of whether they were in the control or study group; and (5) Evaluate perinatal outcomes of pregnant people with obesity who gained fewer than 10 pounds during pregnancy compared to those who gained 10 pounds or more, irrespective of whether they were in the control or study group. | 232 | 12-28 weeks through delivery | prenatal | postpartum | Overweight/obesity | Diet | Reduced GWG; lower rates of HDP | Clinic | No | Caucasian, Indian, Latina, African American | yes | obese |
| Trout 2016 | Macronutrient composition or social determinants? Impact on infant outcomes with gestational diabetes mellitus | Evaluate at 2 socioeconomically diverse sites: 1) effects of a maternal carbohydrate-restricted diet vs usual pregnancy diet on maternal outcomes of blood glucose, weight gain, common medical comorbidities, and incidence of medical procedures and 2) effects of a maternal carbohydrate restricted diet versus usual pregnancy diet on infant outcomes of birthweight, macrosomia, and adverse perinatal events | 68 | 24-28 weeks (GDM diagnosis) up to 35 weeks through delivery | prenatal | prenatal | Gestational diabetes | Diet | Null | Clinic | No | Race: White non-Hispanic, Black, Asian, Hispanic | yes | none |
| Twedt 2023 | Sleep intervention and glycemic control in gestational diabetes mellitus: a feasibility study | Determine the feasibility of a sleep education program targeted to pregnant women with GDM | 70 | GDM diagnosis through 35 weeks | prenatal | prenatal | Gestational diabetes | Other: sleep | Feasibility | Clinic | No | Only race provided: White, Black, Asian, Other | no | none |
| VanHorn 2018 | Dietary Approaches to Stop Hypertension Diet and Activity to Limit Gestational Weight: maternal Offspring Metabolics Family Intervention Trial, a Technology Enhanced Randomized Trial | Investigate whether a calorie-controlled, Dietary Approach to Stop Hypertension (DASH)–type diet and lifestyle intervention guided by a Registered Dietitian Nutritionist (RDN) coach using commercially available weight-loss technology could safely be applied to limit GWG and improve diet quality and physical activity. | 281 | 20 weeks | prenatal | prenatal | Overweight/obesity | Diet; Physical activity | Reduced GWG | Clinic | Yes | Ethnicity (Hispanic/non-Hispanic) is separated, races reported are White, Black or African American, and Other | yes | overweight or obese |
| Vesco 2014 | Efficacy of a group-based dietary intervention for limiting gestational weight gain among obese women: a randomized trial | Determine whether the weight management model often used in nonpregnant adults (weekly, group-based weight management intervention focused on diet and behavior change) would be effective among pregnant people with obesity for limiting GWG and reducing proportion of LGA infants | 114 | 8-21 weeks through delivery | prenatal | postpartum | Overweight/obesity | Diet; Physical activity | Reduced GWG; reduced PPWR | Clinic | No | Only White reported | yes | obese |
| Yeo 2008 | A comparison of walking versus stretching exercises to reduce the incidence of preeclampsia: a randomized clinical trial | Compare a walking exercise to a stretching program during pregnancy in high-risk pregnant people who were sedentary and had previously experienced preeclampsia. | 79 | ~18 weeks through delivery | prenatal | prenatal | Hypertensive disorders of pregnancy | Physical activity | increased transferrin levels | Clinic | No | Only non-Hispanic White reported, everyone else is Other | yes | none |
